# Quantitative MRI susceptibility mapping reveals cortical signatures of changes in iron, calcium and zinc in malformations of cortical development in children with drug-resistant epilepsy

**DOI:** 10.1101/2020.09.15.20157123

**Authors:** Sara Lorio, Jan Sedlacik, Po-Wah So, Harold G. Parkes, Roxana Gunny, Ulrike Loebel, Yao-Feng Li, Emma Dixon, Sophie Adler, J. Helen Cross, Torsten Baldeweg, Thomas S. Jacques, Karin Shmueli, David W Carmichael

## Abstract

**Objective:** Malformations of cortical development (MCD), including focal cortical dysplasia (FCD), are the most common cause of drug-resistant focal epilepsy in children. Histopathological lesion characterisation demonstrates abnormal cell types and lamination, alterations in myelin (typically co-localised with iron), and sometimes calcification. Quantitative susceptibility mapping (QSM) is an emerging MRI technique that measures tissue magnetic susceptibility (χ) reflecting it’s mineral composition.

In a retrospective observational study, QSM was investigated abnormal tissue composition group of children with focal epilepsy with comparison to effective transverse relaxation rate (R2*) and Synchrotron radiation X-ray fluorescence (SRXRF) elemental maps. Our primary hypothesis was that reductions in χ would be found in FCD lesions, resulting from alterations in their iron and calcium content. We also evaluated deep grey matter nuclei for changes in χ with age.

**Methods:** QSM and R2* maps were calculated for 40 paediatric patients with suspected FCD (18 histologically confirmed) and 17 age-matched controls.

Patients sub-groups were defined based on concordant electro-clinical or histopathology data. Quantitative investigation of QSM and R2* were performed within lesions, using a surface-based approach with comparison to homologous regions, and globally within deep brain regions using a voxel-based approach with regional values modelled with age and epilepsy as covariates.

Synchrotron radiation X-ray fluorescence (SRXRF) was performed on brain tissue resected from 4 patients to map changes in iron, calcium and zinc and relate them to MRI parameters.

**Results:** Compared to fluid-attenuated inversion recovery (FLAIR) or T1Lweighted imaging, QSM improved lesion conspicuity in 5% of patients.

In patients with well-localised and confirmed FCDIIb lesions, quantitative profiling demonstrated decreased χ, but not R2*, across cortical depth with respect to the homologous regions. Contra-lateral homologous regions additionally exhibited increased χ at 2-3mm cortical depth that was absent in lesions. The iron decrease measured by the SRXRF in FCDIIb lesions was in agreement with myelin reduction observed by Luxol Fast Blue histochemical staining.

SRXRF analysis in two FCDIIb tissue samples showed increased zinc and calcium, and decreased iron in the brain region exhibiting low χ and high R2*. QSM revealed expected age-related changes in the striatum nuclei, substantia nigra, sub-thalamic and red nucleus, but these changes were not altered in epilepsy.

**Conclusion:** QSM non-invasively revealed cortical/sub-cortical tissue alterations in MCD lesions and in particular that χ changes in FCDIIb lesions were consistent with reduced iron, co-localised with low myelin and increased calcium and zinc content. Theses findings suggests that the measurements of cortical χ measurements could be used to detect and delineate epilepsy lesions.

## Introduction

Malformations of cortical development (MCD), and in particular focal cortical dysplasia (FCD), are the most common cause of drug-resistant focal epilepsy in children (Harvey et al., 2008; Blumcke et al., 2017). The main neuro-pathological features characterising FCD lesions are disrupted cortical lamination, present in FCDI, and the presence of dysmorphic neurons (in FCDIIa sub-type) and balloon cells (in FCDIIb sub-type) in the cortex and white matter (Blümcke et al., 2011), which could be the result of a developmental abnormality involving several phases of corticogenesis (Zucca et al., 2015).

Previous studies have shown that FCDIIb lesions exhibit decreased myelin (Blümcke et al., 2011; Blumcke et al., 2017), although it remains unclear the extent to which they have altered levels of calcium and iron (Aggarwal et al., 2018). Tissue property changes underlying epilepsy have been widely investigated using animal models that have shown calcification and oxidative stress markers in epileptogenic regions (Gayoso et al., 2003; Löscher, 2011; Zimmer et al., 2019). Iron is involved in many fundamental neural processes such as myelin production, oxygen transportation as well as synthesis and metabolism of neurotransmitters (Ward et al., 2014). Myelin plays an important role in the development and function of neocortex, which is characterised by a layered structure with region-specific myeloarchitecture and cytoarchitecture (Fatterpekar et al., 2002; Waehnert et al., 2014; Dinse et al., 2015; Timmler and Simons, 2019). Although myeloarchitecture varies across cortical regions, there is a consistent pattern of high myelination in deep cortical layers compared to superficial ones that is expected to be disrupted in the presence of altered laminar structure.

Calcification and iron accumulation occurs in numerous neurological disorders with possible neurotoxic consequences such as oxidative damage and cell death (Casanova and Araque, 2003). Whilst the mechanisms underlying mineral and metal accumulation are not yet fully understood, measuring altered chemical composition in FCD lesions non-invasively may provide an important biomarker for local pathological processes. Iron accumulation in deep brain regions characterises brain ageing (HALLGREN and SOURANDER, 1958; Ordidge et al., 1994; Aquino et al., 2009; Yao et al., 2009; Li et al., 2014; Acosta-Cabronero et al., 2016; Ashraf et al., 2018; Ghassaban et al., 2019) and drug-resistant epilepsy could potentially hasten this process.

Currently, the non-invasive techniques employed to assess the presence of metal and mineral content in the brain are measurements of the effective transverse relaxation rate (R2*=1/T2*) and more recently, quantitative susceptibility mapping (QSM). R2* is sensitive to both macroscopic and microscopic magnetic field inhomogeneities, the latter may occur due to the presence of iron or myelin in tissue (Ordidge et al., 1994; Yao et al., 2009; Ning et al., 2014; Homayoon et al., 2019; Tabelow et al., 2019). QSM provides a quantitative and local anatomical contrast by estimating the magnetic susceptibility (χ) using the MRI signal phase (Haacke et al., 2015, p. 201, Liu et al., 2015*a*; Deistung et al., 2017). Typically, in the human brain a strong χ image contrast is obtained in structures containing paramagnetic substances, such as iron-rich deep brain nuclei showing positive χ values relative to the surrounding tissue (Bilgic et al., 2012; Zheng et al., 2013), and in regions with diamagnetic substances, such as highly myelinated white matter fibres and calcifications exhibiting negative χ values (Li et al., 2011; Langkammer et al., 2012). Therefore, we would expect QSM to be sensitive to the calcium alterations in lesions and to the disruption of the myeloarchitecture in the cortex.

Both χ and R2* values are sensitive to age-related changes of iron in deep grey matter nuclei (Yao et al., 2009), but χ can distinguish better then R2* between sources of paramagnetic and diamagnetic substances. For this reason QSM has been used clinically to differentiate between haemorrhages and calcifications (Klohs et al., 2011; Liu et al., 2012; Lotfipour et al., 2012; Wisnieff et al., 2015;Deistung et al., 2013*b*; Chen et al., 2014). However, the only application of QSM to focal epilepsy so far has been limited to animal models (Aggarwal et al., 2018).

The aim of this study is the first non-invasive characterisation of abnormal tissue mineral composition using QSM in a group of children with focal epilepsy. Therefore QSM mapping was performed in patients with MRI visible lesions consistent with malformations of cortical development. Our primary hypothesis was that *local* reductions in χ would be found in lesions due to underlying alterations in iron or calcium content. This hypothesis was investigated in a group of subjects by evaluating changes radiologically and quantitatively. We investigated changes in the expected laminar distribution of cortical iron at the group level using a surface-based approach, sampling at increasing depths from the pial surface within lesions and homologous regions. Further, in a small group changes were compared to synchrotron radiation X-ray fluorescence (SRXRF) elemental measurements of iron, calcium and zinc content (Coulter, 2000; Zheng et al., 2013; Carver et al., 2016).

Our secondary hypothesis was that there could be global brain iron changes in the deep grey matter nuclei in children with poorly controlled epilepsy, reflecting the patho-physiological changes within extended epileptic networks (Centeno et al., 2014; He et al., 2017). In order to test this, we measured changes in with age in the deep brain nuclei of healthy control subjects and assessed if this relationship was different in subjects with focal epilepsy. The same analysis was repeated with the R2* measurements.

## Methods

### Participants

A retrospective cohort of 40 patients (mean age = 8.8 ± 5 years, range=2-21 years, female=16) was identified for this research study from all those undergoing assessment for epilepsy surgery at Great Ormond Street Hospital (GOSH), following approval by the national research ethics service. Patients were included if they had a diagnosis of suspected FCD based on radiological and electro-clinical reports. All patients underwent video-telemetry–EEG to document seizures and 3T MRI at GOSH with the standard epilepsy imaging protocol. Patients were excluded if MRI scans showed severe motion artefacts (i.e. indistinguishable adjacent gyri due to motion or severe ringing), they were younger than 2 years of age, or they were without the full MRI protocol described in the following section.

From the full cohort of 40 patients, a sub-group was defined with well-localised lesions. The criteria for including patients in the well-localised group were as follows: having resection inclusive of the MRI defined lesion with histopathological confirmation, or presenting with an MRI-visible lesion concordant with the electro-clinical data.

The cohort of healthy controls is described in the Supplementary material together with the image acquisition protocol used to scan them.

### MRI protocol

All patients were scanned on a 3T whole-body MRI system (Magnetom Prisma, Siemens Medical Systems, Germany), using a 20-channel receive head coil and body coil for transmission. The clinical protocol included 3D T1-weighted (T1w) images acquired using magnetisation prepared rapid gradient echo (MPRAGE) (repetition time (TR) = 2300ms, echo time (TE) = 2.74ms, matrix size 256×208×256, flip angle=8°, voxel size=1×1×1mm^3^), 3D FLAIR (TR=4000ms, TE=395ms, inversion time (TI)=1800ms, matrix size = 306×192×384, flip angle=120°, voxel size=0.65×1×0.65mm^3^), and three-dimensional T2*-weighted gradient echo images used to compute R2* and QSM (TR= 38ms, flip angle=15deg). The T2*-weighted images were acquired with a monopolar multi-echo acquisition at seven equidistant TE between 3 and 27ms, resolution=1.15×1.15×1.15mm^3^, matrix size 156×192×144, and approximately 6 min acquisition time. Parallel imaging was used along the in-lane phase-encoding (PE) direction (acceleration factor 2 GRAPPA reconstruction), 6/8 partial Fourier was applied in the partition direction. This sequence was also used for computing susceptibility-weighted images (SWI) with minimum intensity projection (mIP). The SWI and mIP were included in the clinical protocol in order to provide state-of-the-art evaluation of vessel micro-bleeds and vascular abnormalities that are relatively common in this cohort(Craven et al., 2012).

### Calculation of QSM and R2* maps

To reconstruct the QSM map, an initial brain mask was calculated based on the first TE magnitude image using the Brain Extraction Tool (BET) (Smith, 2002) from FSL6.0 (www.fmrib.ox.ac.uk/fsl). The initial brain mask was iteratively eroded in border areas with too high (>97.5%) or too low (<2.5%) phase values in the local phase map, as these phase values were considered artefacts. The iteration stopped when the standard deviation of the phase values of the border of the mask was below 1.2 times of the standard deviation of the phase values within the mask. The iteration also stopped, if the local mask area was smaller than 70% of the initial mask.

The processing pipeline for QSM consisted of: non-linear fitting of the complex GRE signal over multiple echoes(Liu et al., 2015*a*); spatial phase unwrapping using Laplacian kernels (Schweser et al., 2013), background field removal using the Laplacian boundary value method (Zhou et al., 2014), local field-to-susceptibility inversion using direct Tikhonov regularization (Kressler et al., 2010), and correction for the susceptibility underestimation (Schweser et al., 2013). For the Tikhonov-based regularization, the optimal regularization parameter was calculated as the average across 10 representative subjects of the individual optimal regularization parameters calculated using the L-curve method (Hansen and O’Leary, 1993).

R2* maps were computed using a voxel-wise linear fit of the logarithm of the magnitude image over the TE of the T2*-weighted dataset.

### Visual assessment

To evaluate the lesion visibility on the QSM and R2* and compare it to the clinical images, two neuro-radiologists (RG, UL) were presented with coregistered T1w, FLAIR, QSM and R2* images. Following standard radiological practice, they were allowed to compare between the different contrasts in the same patient; this allowed them to visually assess lesion location and relative conspicuity in the different image types. A lesion visibility score from 1 to 4 (1=not visible, 2=subtle, 3=visible, 4=clearly visible) was assigned to each image type for each patient. The observers were not blinded to the radiological and EEG reports (i.e.: localisation of the lesion) because the aim was to derive relative conspicuity *not* test lesion detection performance. Intensity windowing was individually adapted to gain optimal contrast. The neuro-radiologists assessed the images independently and so were blinded to each other’s ratings.

To compare lesion conspicuity scores between T1w, FLAIR, R2* and QSM images across patients, we applied Friedman test to the mean score of the two radiologists computed for each map. Then we performed a multiple comparison test between the ranking means provided by the Friedman test for each group of images. We set statistical significance at p<0.05 after applying the Bonferroni correction for multiple comparisons.

### Image processing

#### Sampling of cortex and subcortical white matter

Figure 1 shows the workflow for the sampling of the QSM and R2* data. *FreeSurfer* software v5.3 (Dale et al., 1999; Fischl and Dale, 2000) was used to co-register the QSM, R2* maps and FLAIR to the T1w image, and to reconstruct cortical and subcortical surfaces. Both FLAIR and T1w images were employed to generate the surface reconstruction. The image processing steps included skull-stripping (Ségonne et al., 2004), intensity non-uniformity correction (Sled et al., 1998), automated Talairach transformation, extraction of the deep GM structures (including hippocampus, amygdala, caudate, putamen, ventricles) (Fischl et al., 2004), and intensity normalisation (Sled et al., 1998). Finally the images were tessellated and deformed to create accurate smooth mesh representations of the pial surface(Fischl and Dale, 2000; Ségonne et al., 2007).

**Figure 1:**
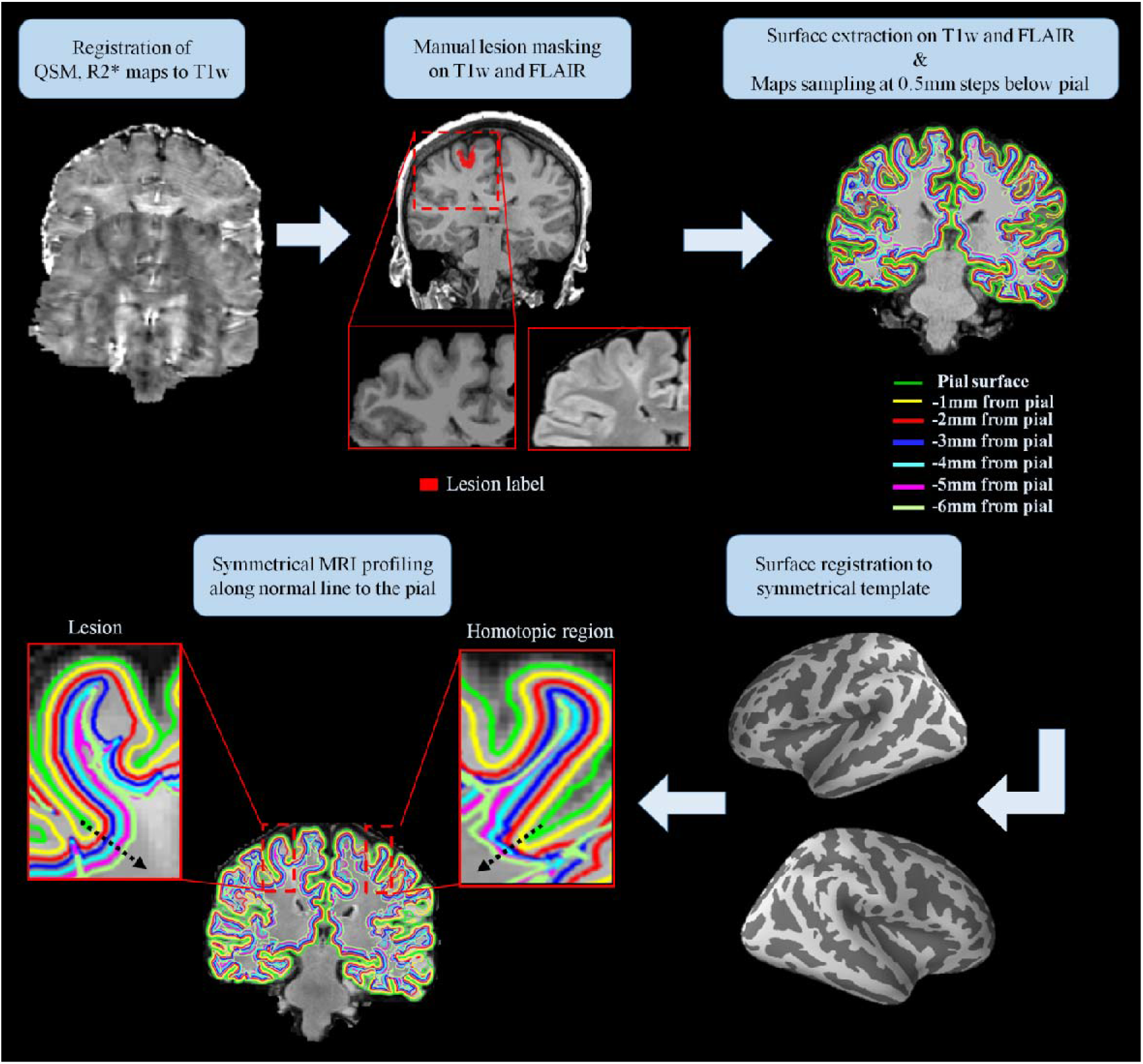
Maps sampling workflow. First the QSM and R2* maps were coregistered to the T1-weighted (T1w) data. Manual lesion masking and surface extraction at increasing depths were performed on T1w and FLAIR images. The QSM and R2* maps were projected onto the surfaces and sampled at several depths from the pial surface. The sampled surfaces and the lesion masks were then spatially registered to a symmetric template, allowing the symmetrical MRI profiling of the QSM and R2* maps along the normal (black arrow) to the pial surface for each lesion and the corresponding homotopic region.

To examine the cortical and subcortical signal of the QSM and R2* maps while minimising GM -WM misclassification errors, the QSM and R2* maps were sampled along surfaces extracted at different depths from the pial surface down to 6mm at steps of 0.5mm. This avoided the potential confounds related to identifying the white/grey matter border in FCD where often no clear border exists, and which can cause the cortical thickness to be mis-estimated. Then sampled QSM and R2* maps were smoothed using a 10mm FWHM Gaussian kernel, and they were registered to an average symmetric space having an identical number of vertices for each hemisphere (Greve et al., 2013). This allowed us to analyse the χ and R2* profile changes between homologous regions, guided by a straight line orthogonal to the cortical surface that provided vertex correspondence across surfaces (Polimeni et al., 2010). While this does not fully account for variable cortical thickness, it allows to use homologous vertexes as internal controls where the contralateral region would be expected to have comparable thickness in the absence of pathology.

### Lesion masking

FCD lesions were identified on T1w and FLAIR images by an experienced paediatric neuro-radiologist. 3D binary masks were manually delineated for the 40 patients. The lesion masks were first registered onto the *FreeSurfer* surface reconstructions and then to the symmetric template as shown on Figure 1. This procedure provided a mask for the lesion and one for the homologous healthy tissue.

### Segmentation of deep brain regions

In order to perform age regression models using QSM and R2* maps, deep brain nuclei characterised by high iron content, were delineated using a binary mask (Lorio et al., 2016). The regions analysed were the globus pallidus, putamen, caudate, thalamus, substantia nigra, subthalamic nucleus, red nucleus and cerebellar dentate. The masks were warped from the MNI space to the subject’s native space using the deformation fields estimated with SPM12 (Wellcome Trust Centre for Neuroimaging, London, UK; http://www.fil.ion.ucl.ac.uk/spm) and Matlab 9.5 (Mathworks, Sherborn, MA, USA). The deformation fields were computed by performing tissue classification on the T1w images using the “segmentation” approach in SPM12 (Ashburner and Friston, 2005). The suspected FCD patients did not exhibit any lesions in the deep brain nuclei.

### Statistical analysis

#### Lesion profiling

MRI profiles for the QSM and R2* maps were obtained by averaging the values within each patient’s masks for the lesion and homologous region, separately, along cortical and subcortical surfaces. To investigate signal changes in FCD lesions, QSM and R2* profiles were statistically compared between lesion and homologous regions for the 40 patients. The same analysis was repeated within the sub-group of patients with FCD IIb, and within the well-localised sub-group.

We used a paired t-test to evaluate χ and R2* differences at various sampling depths. Correction for multiple comparisons was applied using false discovery rate (FDR) at q<0.05. Data normality, required for the t-test, was assessed using the Shapiro-Wilk test run on the MRI profiles of the lesions and homologous regions.

#### Age effect in deep brain nuclei

To test for a linear correlation of χ and R2* values with age, a multiple regression model was applied to both the whole cohort of patients and controls. The same analysis was repeated on the controls and the following sub-groups: 15 patients with FCDIIb and 25 patients with well-localised lesions.

For each deep brain structure the mean χ and R2* was modelled using age, disease duration and disease presence as independent variables. The regression was implemented using the ordinary least square method embedded in IBM SPSS v25. In order to test for the interaction between age and disease presence on the χ and R2* respectively, a hierarchical multiple regression was applied. This model assessed the increase in variance explained by the addition of the interaction term between age and disease presence to the main effects model.

### SRXRF elemental mapping and analysis

To identify and quantify the element driving QSM changes in the FCD lesions, we used SRXRF to simultaneously quantify and spatially evaluate elements in the brain tissue resected during surgery. One subject without diagnostic findings and three with different pathologies were selected on the basis of having clear visually apparent changes in QSM maps within the lesion and recovery of sufficient amounts of tissue from surgical resection. In order to allow the registration of the specimen with the MRI maps, we selected tissue samples containing white and grey matter with partially preserved anatomical borders.

Each brain specimen was placed in formalin shortly after surgery and processed for embedding in wax for histological examination using standard diagnostic testing that included haematoxylin and eosin (H&E) staining, Luxol Fast Blue/Cresyl Violet staining (LFB) and immunohistochemistry as required for diagnosis. The detailed description of staining techniques applied is reported in the Supplementary material.

Microtomy was performed to produce 7μm thick formalin fixed paraffin embedded sections that were mounted onto 4μm thick Ultralene film (Spex Sample-Prep, NJ, USA) secured to a customized holder for SRXRF.

SRXRF of the brain tissue sections was performed on the I18 beam-line at the Diamond Light Source synchrotron radiation facility (Didcot, UK) as described by Walker and colleagues (Walker et al., 2016). Briefly, the beam energy was tuned to 11keV and focused to 100 × 100 μm (determining resolution) and the tissue samples scanned in a raster manner. The samples were mounted at a 45° angle with respect to the incoming X-ray beam and the detector to minimize scatter contribution. The raw data consist of full energy dispersive spectra for each sample point exposed to the beam. The spectra were subsequently fitted using PyMca (Solé et al., 2007) and the areas of the characteristic peaks for iron, zinc and calcium evaluated. Quantification was performed by measuring a reference metal film (AXO, GmbH, Dresden, Germany) to estimate photon flux on the samples (Walker et al., 2016). SRXRF elemental maps of pixel-by-pixel elemental iron, zinc and calcium concentrations (parts per million, ppm) were manually aligned to the corresponding histology staining using ImageJ. Regions of interest (ROIs) were manually drawn on the SRXRF maps, and were used to quantify iron, zinc and calcium (mean ± standard deviation) using ImageJ (https://imagej.nih.gov/ij/).

## Results

### Patients’ clinical information

The patients’ demographics and clinical information can be found in Table 1. A total of 25/40 patients underwent surgery: 18 were histologically diagnosed with FCD (15 FCDIIb, 3 FCDIIa, 16 had Engel outcome ≥II), one had laser ablation, therefore the histological examination was not possible, one had polymicrogyria, two had glioneuronal tumours, three were diagnosed with hippocampal sclerosis (HS) and had standard temporal lobe resection involving the anterior temporal lobe, histopathological analysis did not exhibit any specific diagnostic pathology outside the hippocampus in these cases (Table 1). Seizure freedom was achieved in 19/25 cases at a mean of 1.5 years after surgery.

**Table 1:** Patient demographics. Age is presented in a range of years <5, 6-10,11-15, 16+, while seizure onset is presented in years (y), months (m) or weeks (w). The lesion location is reported according to the MRI-based radiological report. Electrophysiological clinical information is provided based on the EEG seizure localisation. If multiple lesion locations were reported in patients who underwent surgery, all locations were resected. Surgery outcome for the patients who underwent surgery is classified using the Engel post-operative surgical outcome according to Engel classification (Engel, 1993), Ia = completely seizure free, III = worthwhile improvement, IV = no worthwhile improvement. Abbreviations: m=male, f=female, L=left, R=right, fr= frontal pole, ant= anterior, inf= inferior, occ=occipital pole, par=parietal pole, post=posterior, sup=superior, temp=temporal lobe, FCD=focal cortical dysplasia, HS=hippocampal sclerosis, n=no, y=yes.

| Patient | Gender<br>(m=0) | Age<br>group<br>(y) | Disease<br>onset | MRI |  | EEG |  | Histology | Surgery | Engel |
| --- | --- | --- | --- | --- | --- | --- | --- | --- | --- | --- |
|  |  |  |  | site | radiological report | localising | site |  |  |  |
| 1 | 0 | <5 | 22m | R mesial par-occ | focal signal abnormality | y | R post | Glioneuronal tumour | y | Ia |
| 2 | 0 | 16+ | 3y | R inf fr gyrus, fr operculum | focal signal abnormality | y | R fr | FCD IIb | y | Ia |
| 3 | 0 | <5 | 1y | L fr, sup longitudinal sulcus | subtle lesion | n |  | NA | n |  |
| 4 | 1 | 6-10 | 2y7m | L par operculum | abnormal cortical folding, blurring | y | L side predominance | FCD IIb | y | III/IV |
| 5 | 0 | <5 | 3y | R post fr, ant motor strip | blurring | y | R post temp lobe | FCD IIa | y | Ia |
| 6 | 0 | 11-15 | 2m | L paracentral lobule | focal signal abnormality | y | L paracentral lobule | FCD IIb | y | Ia |
| 7 | 1 | 16+ | 6y | L sup fr gyrus | focal signal abnormality | y | L front area | No abnormality | y | Ia |
|  |  |  |  | L mesial temp | hippocampal sclerosis |  |  | HS (ILAE 3) |  |  |
| 8 | 0 | 6-10 | 11m | L sup fr gyrus | focal signal abnormality | y | L frontotemporal | FCD IIb | y | II |
| 9 | 0 | 6-10 | 3d | R fr, ant temp | smaller hemisphere, diffuse blurring | y | R fr | FCD IIb | y | II |
| 10 | 0 | 6-10 | 8y | R post temp | focal signal abnormality, trans-mantle sign | n |  | NA | n |  |
| 11 | 0 | <5 | 3y | R temp | focal signal abnormality | n | diffuse | NA | n |  |
| 12 | 0 | 11-15 | 3m | R par-occ | blurring | n | multifocal | NA | n |  |
| 13 | 1 | 16+ | 1y | L post cingulate gyrus | focal signal abnormality | n |  | No abnormality | y | Ia |
|  |  |  |  | L mesial temp | hippocampal sclerosis |  |  | HS (ILAE 1) |  |  |
| 14 | 1 | 6-10 | 9m | R middle, inf fr, sup par gyri | focal signal abnormality | n | diffuse | NA | n |  |
| 15 | 0 | 16+ | 1y | L inf temp sulcus | focal signal abnormality | y | temp and fr both sides | FCD IIb | y | Ia |
| 16 | 1 | 11-15 | 3w | L temp | white matter signal change | y | L paracentral | NA | n |  |
| 17 | 0 | 16+ | 9y | L fr gyrus | focal signal abnormality | n |  | NA | n |  |
| 18 | 0 | 11-15 | 2y | L sup temp gyrus | focal signal abnormality | y | L temp | FCD IIb | y | Ia |
| 19 | 0 | 16+ | 12y | L occ | subtle difference in grey-white matter differentiation | n | multifocal fronto-parietal | NA | n |  |
| 20 | 0 | 11-15 | 2m | L paracentral lobule | focal signal abnormality | y | L paracentral | FCD IIb | y | Ia |
| 21 | 0 | <5 | 2m | L occ | focal signal abnormality | y | L posterior quad | NA | n |  |
| 22 | 1 | 6-10 | 6y | L temp | focal signal abnormality | y | L post temp lobe | glioneuronal tumour | y | Ia |
| 23 | 0 | <5 | 9m | L fr | focal signal abnormality | y | L fr | NA | n |  |
| 24 | 1 | 6-10 | 23m | L fr | blurring | y | R fr onset and L fronto-temporal | FCD IIa | y | III/IV |
| 25 | 1 | <5 | 8w | L sup temp gyrus | focal signal abnormality | y | Left temporal | FCD IIb | y | Ia |
| 26 | 1 | <5 | 13m | R middle temp gyrus | focal signal abnormality | y | Right temporal posterior | FCD IIb | y | Ia |
| 27 | 0 | 11-15 | 1.5y | R temp | hippocampal sclerosis |  |  | HS (ILAE 1) |  |  |
|  |  |  |  | R middle temp gyrus | focal signal abnormality | y | R temp lobe | No abnormality | y | III/IV |
| 28 | 0 | 11-15 | 6y | R fr and insular lobes | focal signal abnormality | y | R fr lobe | Polymicrogyria | y | Ia |
| 29 | 0 | 6-10 | 7m | R int and mesial fr lobe | blurring | n |  | NA | n |  |
| 30 | 1 | 11-15 | 11y | L precentral gyrus | focal signal abnormality | n |  | NA | n |  |
| 31 | 0 | 6-10 | 3m | R par and occ | blurring | n | Multifocal | NA | n |  |
| 32 | 0 | 6-10 | 9w | L post insular region | white matter signal change | y | (Stereoeeg) L insula | NA due to laser ablation | y | Ia |
| 33 | 0 | 6-10 | 7y | R sup and middle fr gyri | focal signal abnormality | n |  | NA | n |  |
| 34 | 1 | 11-15 | 14y | L temp | subcortical lesion | y | L ant cortical | NA | n |  |
| 35 | 1 | <5 | 8m | R fr | focal signal abnormality | y | R fr | FCD IIa | y | Ia |
| 36 | 1 | 6-10 | 1y | L fr | focal signal abnormality | y | L fr | FCD IIb | y | II |
| 37 | 1 | <5 | 6.5m | R occ | focal signal abnormality | y | R post-temp | FCD IIb | y | Ia |
| 38 | 1 | 6-10 | 1y | R fr | focal signal abnormality | y | R ant | FCD IIb | y | Ia |
| 39 | 0 | <5 | 1y | L temp | focal signal abnormality | y | L temp | FCD IIb | y | Ia |
| 40 | 0 | 6-10 | 2y | R fr | focal signal abnormality | y | R mostly fr | FCD IIb | y | Ia |

We identified two sub-groups with n≥10 patients: the one with 15 FCDIIb patients and the group of patients with well-localised lesions. The latter was composed of 25 patients: 21 that received a histopathological diagnosis following epilepsy surgery (15 FCDIIb, 3 FCDIIa, 1 polymicrogyria, 2 Glioneuronal tumours), and 4 patients without surgery, having the MRI-visible lesion concordant with the electro-clinical data. Patients diagnosed with HS were not included in the well-localised group due to the lack of diagnostic findings in the resection of the anterior temporal lobe.

### Visual assessment

Examples of patients’ with clear visual alterations in values, spatially concordant with changes visible in FLAIR images, are shown on Figure 2 with comparison to R2* maps.

**Figure 2:**
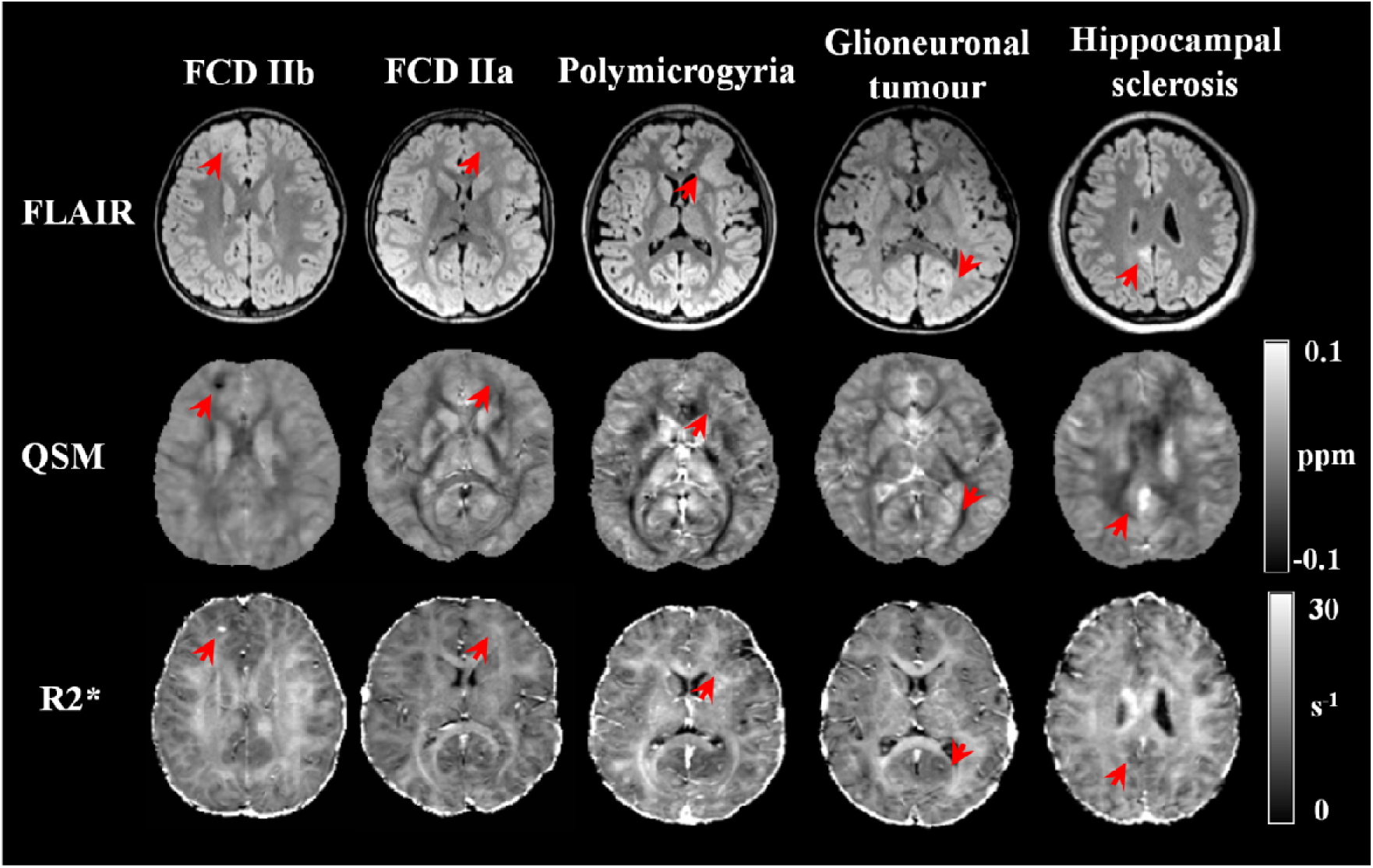
Examples of QSM and R2* maps obtained in the clinical setting for patients with FCDIIb, FCDIIa, polymicrogyria, Glioneuronal tumour and hippocampal sclerosis (HS). The red arrow points toward the lesion identified by radiologists. For the HS patient, the arrow highlights an abnormal region in the posterior cingulate gyrus as identified on the radiological report.

Compared to the best visualisation achieved in either FLAIR or T1w, the lesions conspicuity was visually assessed as being better/equal/worse on the QSM in respectively 2/3/35 individuals and on the R2* in 3/3/34.

The lesion conspicuity was scored as being significantly different across image types (p<0.001). The post-hoc multiple comparison test applied to the Friedman ranking test showed that the mean ranking of FLAIR and T1w was not significantly different, while the mean ranking of R2* and QSM was significantly reduced compared to FLAIR images (p<0.05).

### QSM, R2* maps and SRXRF elemental analysis

The SRXRF analysis was carried out on samples from two FCD IIb patients, a patient with a glioneuronal tumour, and on the temporal lobe tissue resected from of a HS patient. This patient had partial hippocampal and standard temporal lobe resection involving the anterior temporal lobe; however the latter tissue did not show specific diagnostic changes after the histological examination. Therefore, we used the portion of the anterior temporal lobe as control tissue for the experiment.

For the first FCDIIb patient (patient 8), we observed increased zinc and calcium in the tissue compatible with the findings of low χ and high R2* values in the lesion (see Fig. 3). The calcium level in the lesion 15×10^3^ ± 5×10^3^ ppm; while in the neighbouring tissue it was 335 ± 52 ppm. The zinc content in the lesion was 113.7 ± 39.2 ppm, and in the neighbouring tissue it was 13.9 ± 0.9 ppm. Moreover, in the subcortical white matter belonging to the lesion, there was decreased iron (Fe= 16 ± 1 ppm), compared to normal white matter (Fe= 36 ± 10 mg/kg ppm). A visual inspection of the LFB staining showed that the areas exhibiting low myelin were consistent with those with low iron content (see Fig. 3). For the second FCDIIb patient (patient 15) we found decreased iron in the FCD lesion with respect to the neighbouring tissue (see Fig. 4), 19 ± 2ppm and 40 ± 2ppm, respectively. The zinc content in the lesion was 5.2 ± 0.6 ppm; while in the neighbouring tissue it was 9 ± 2.6 ppm. Calcium appeared to be similar in the lesion and surrounding tissues.

**Figure 3:**
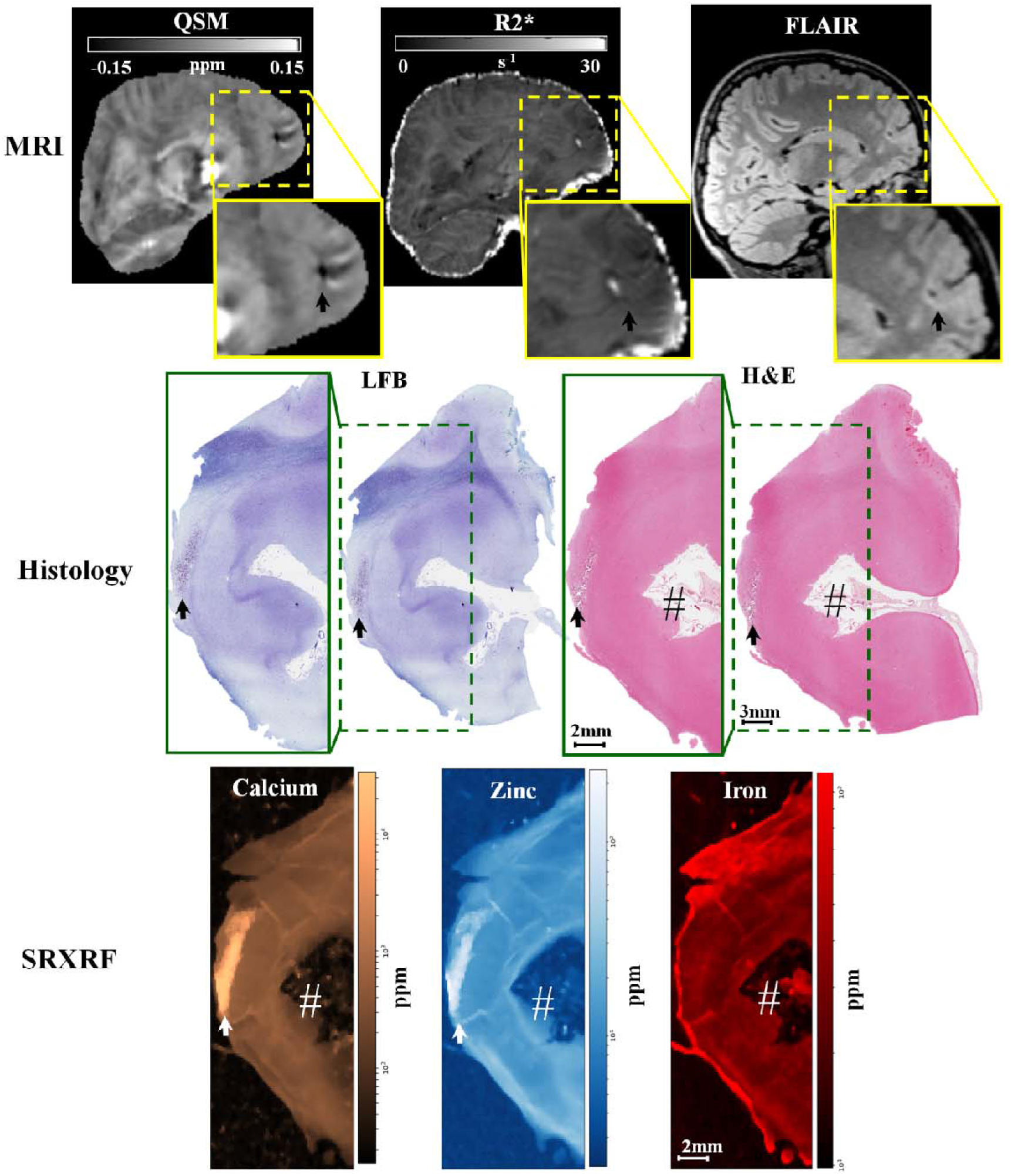
Example 1 of FCDIIb lesion analysed using MRI, histology and synchrotron radiation X-ray fluorescence (SRXRF). In the upper row, the FCD lesion is delineated by hypo-intensity by QSM, hyper-intensity by R2*, and cortical thickening by FLAIR. The middle row shows histological luxol fast blue (LFB) and haemotoxylin-eosin (H&E) staining of sections from the resected tissue. LFB identifies an area with low myelin content (↑) in the white and grey matter, while H&E stains mineral deposits (↑) dark purple. The bottom row shows the SRXRF elemental maps for calcium, zinc and iron. Increased calcium and zinc (↑) are present in the area corresponding to mineral precipitates as assessed by H&E, and to the hypo-intense region (↑) on QSM. Iron content is lower in the subcortical white matter underneath the sulcus (#) compared to that in normal appearing white matter.

In order to assess if the iron changes were related to myelin content variation, we performed additional LFB staining for both patients on a section adjacent to the one used for the SRXRF analysis. A visual inspection of the LFB staining showed that the areas exhibiting low myelin where the ones with low iron content (see Fig. 4).

**Figure 4:**
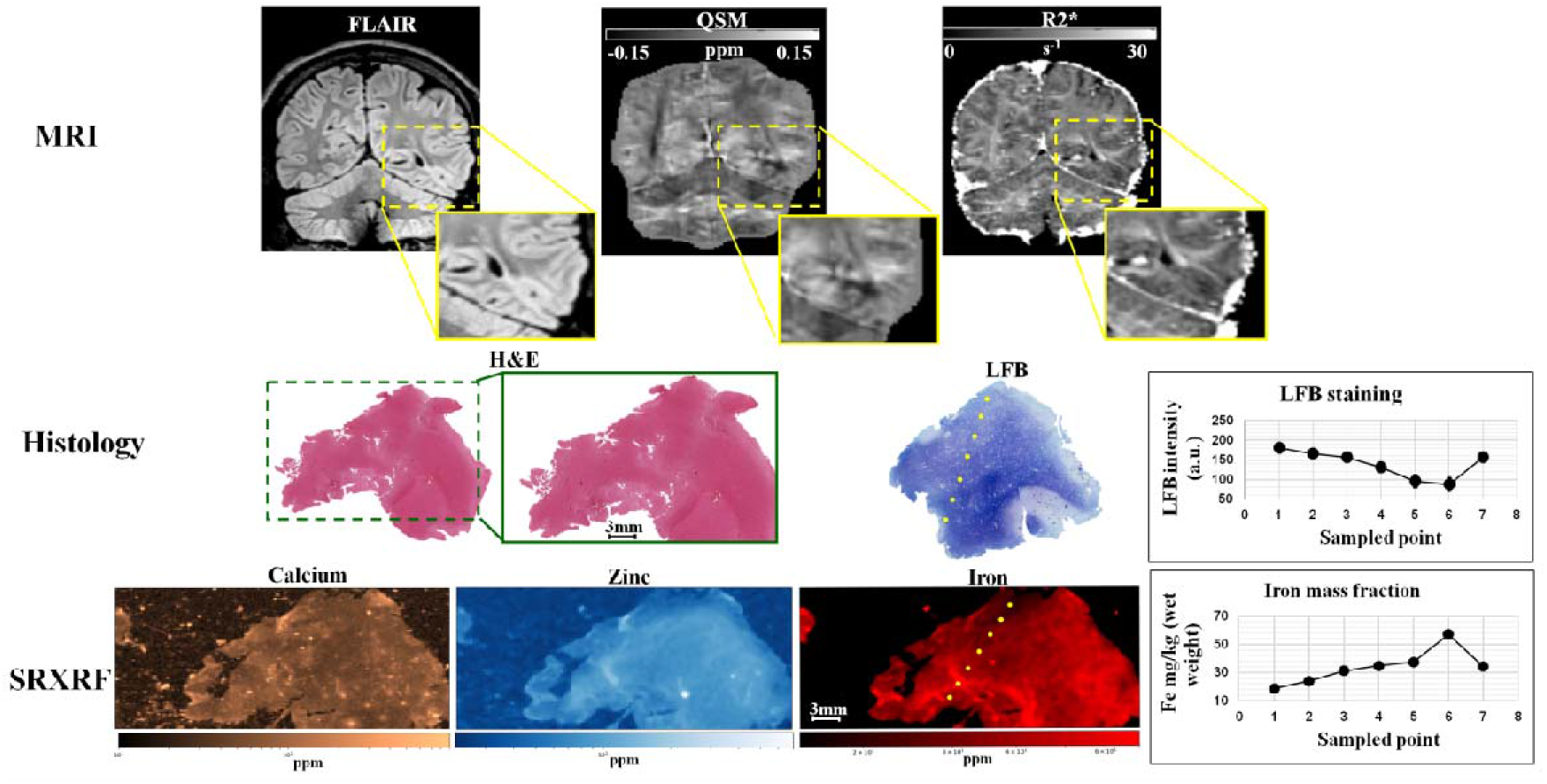
Example 2 of FCDIIb lesion analysed using MRI, histology and synchrotron radiation X-ray fluorescence (SRXRF). In the upper row, the FCD lesion is delineated by cortical thickening on FLAIR and by R2*, and hypo-intensity on QSM. The middle row shows histological haemotoxylin-eosin (H&E) and luxol fast blue (LFB) staining performed on sections of resected tissue. The LFB identifies an area with low myelin content at the top of the dotted line. The LFB intensity profile along the dotted line is plotted on the graph on the right side of the middle row. The bottom row shows the SRXRF elemental maps for calcium, zinc and iron. Low iron content is observed at the top of the dotted line and iron content along the dotted line is plotted on the graph on the right side of the bottom row. Despite the dotted line was arbitrary drawn, the LFB staining intensity and the iron content graphs indicate that myelin increase (decrease in LFB intensity) corresponds to higher iron content.

Additionally, we performed SRXRF analysis on the tumour region from patient 22 diagnosed with glioneuronal tumour. We found: increased calcium (11 x10^3^ ± 7×10^2^ ppm within the tumour compared to adjacent normal appearing grey matter (200 ± 50 ppm), increased zinc (11×10^2^ ± 7×10 ppm) within the tumour compared to normal appearing grey matter (70 ± 50 ppm), increased iron (200 ± 80 ppm) in areas where small blood vessels were present (see Fig. 5). The areas with increased zinc and calcium on the SRXRF maps were consistent with regions showing low χ and high R2* values on the MRI maps.

**Figure 5:**
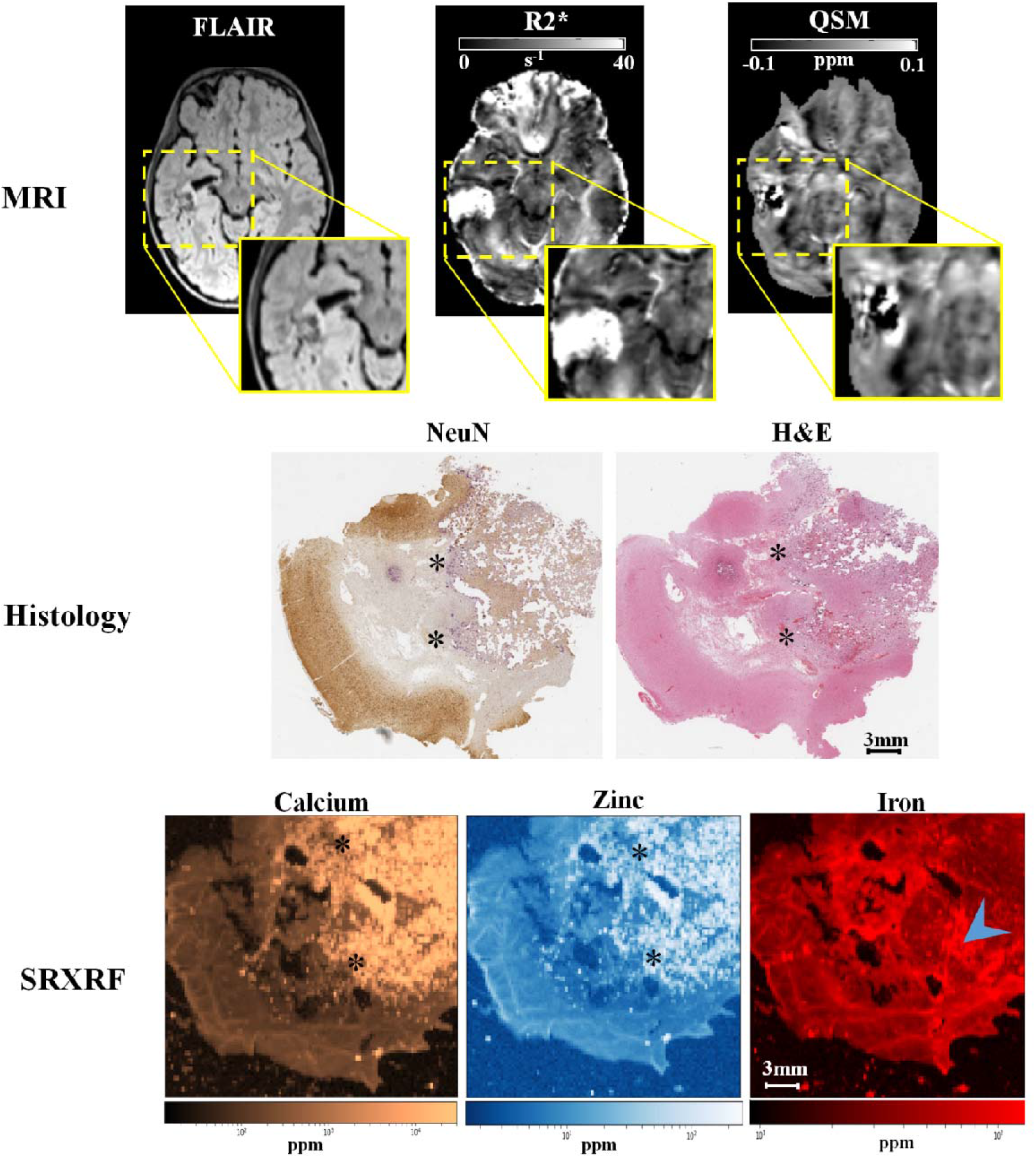
Example of glioneuronal tumour analysed using MRI, histology and synchrotron radiation X-ray fluorescence (SRXRF). In the upper row, the lesion is delineated by abnormal intensity on FLAIR hyper-intensity on R2* and hypo-intensity on QSM. The middle row are optical micrographs of sections from the resected tissue stained with neuraminidase (NeuN) or haemotoxylin-eosin (H&E)section. Both NeuN and H&E identify areas with mineral deposits (*) and stained in dark purple. The bottom row shows the SRXRF elemental maps for calcium, zinc and iron. Increased calcium and zinc (*) are present in those areas identified to be have mineral precipitates by NeuN and H&E staining, and are hypo-intense region on QSM. High iron content is present in the region highlighted by the blue arrow which appears to be blood by H&E staining.

For the control tissue without histopathological abnormalities resected from the anterior temporal lobe, we also quantified calcium, zinc, and iron in both the grey and white matter (Ca WM: 63.33 ± 7.15 ppm Ca GM: 113.23 ± 59.45 ppm; Zn WM: 7.5 ± 0.88 ppm, Zn GM: 12.41 ± 2.8 ppm; Fe WM: 45.82 ± 4.96 ppm, Fe GM: GM: 30.13 ± 1.54 ppm).

### Quantitative lesion profiling

No significant χ differences were found across the whole patient cohort between the lesion profiles and homotopic region ones (see Figure 6). However, when restricting the analysis to well-localised lesions we observed decreased χ values across cortical depths with significant differences at 2-3mm with respect to the homologous regions (p_FDR_<0.05). Similar results were obtained for the sub-group with histologically confirmed FCDIIb lesions, as shown on Figure 6. We did not observe any significant R2* differences between the lesions and homotopic regions in any of the patients groups.

**Figure 6:**
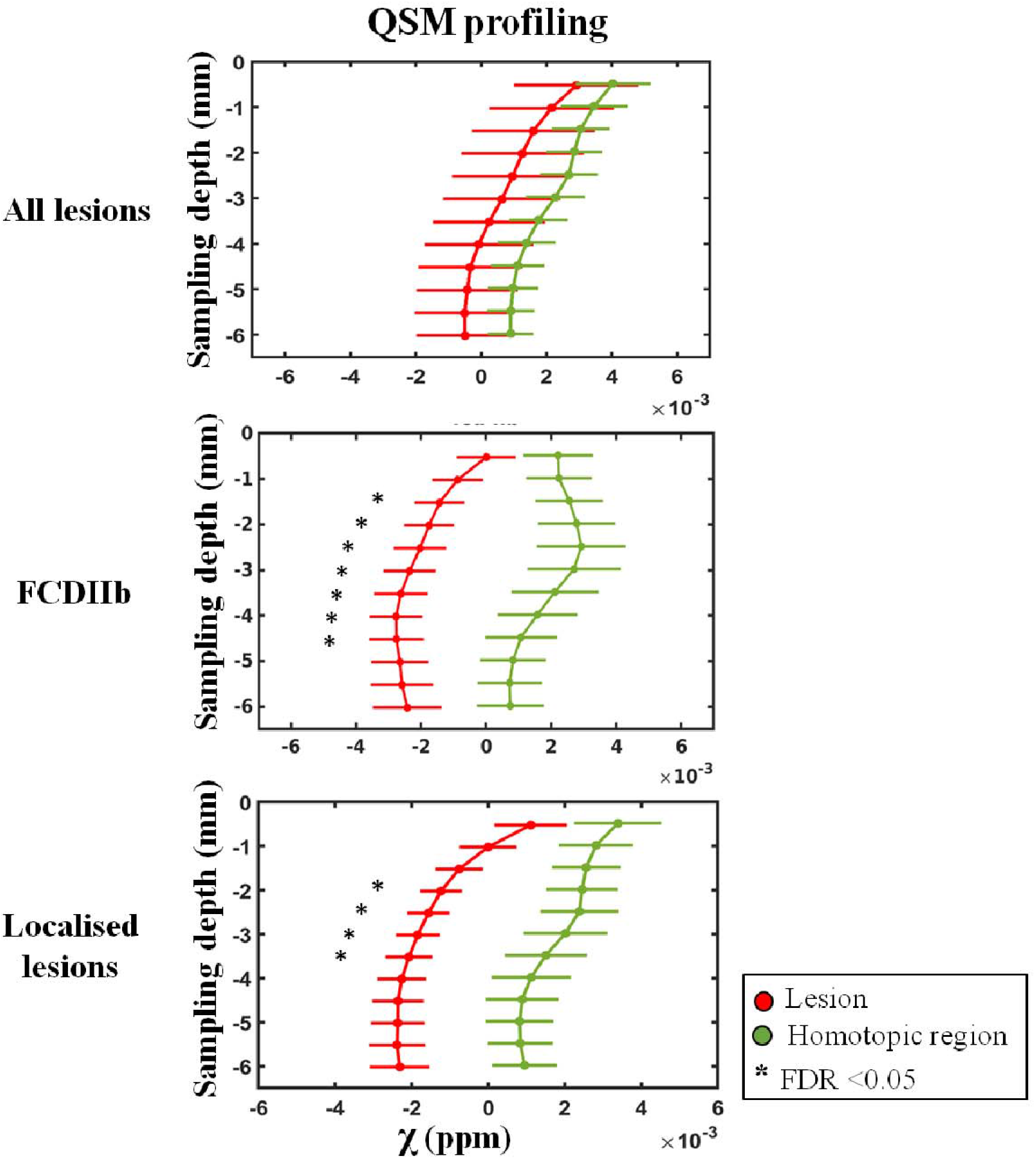
MRI profiling of FCD lesions and homotopic regions. The plots show QSM-based profiling obtained for lesion (red) and homotopic regions (green) for the whole cohort of patients, histology confirmed FCDIIb and well-localised lesions (including histology confirmed lesions and suspected FCD lesions with EEG-based seizure semiology concordant with MRI-based radiology report). The sampling depth is reported as a distance from the pial surface (0mm). Error bars correspond to standard error of the mean. (*) identifies significant profile changes between lesion and homotopic area at false discovery rate (FDR) <0.05.

### Age effect in deep brain nuclei

To determine if age and disease related changes in iron levels in deep brain nuclei could be detected by alterations in χ and R2* values, a multiple linear regression model was used. The mean χ and R2* values for each deep brain nuclei were predicted using regressors for age, disease presence, disease duration and the interaction between disease presence and age in patients and healthy controls. There was linearity of the dependent variables and the predictors, as assessed by partial regression plots and a plot of studentized residuals against the predicted values. There was no evidence of multi-collinearity between independent variables, as evidenced by no tolerance values less than 0.2. The assumption of normality was met, as assessed by a Q-Q Plot.

The multiple regression model significantly (p<0.05) predicted the mean χ in the caudate (left and right), putamen (left and right), pallidum (left and right), substantia nigra (left and right), sub-thalamic nucleus (left and right), red nucleus (left and right), as reported on Table 2. Age was a significant (p<0.05) predictor for the caudate, pallidum and substantia nigra (see Table 2). Disease duration was also significant predictor for the putamen, pallidum and substantia nigra (see Table 2). The regression models applied respectively within the well-localised and FCDIIb patients’ sub-groups together with controls, showed similar age and disease correlation in the same nuclei identified in the whole group analysis.

**Table 2:** Summary of the linear models explaining the mean susceptibility (χ) and R2*. The models were estimated separately for each deep brain structure and each map. The model predictors were the subject’s age, disease presence, disease duration and interaction between the disease effect and age. We report the p-value of each predictor, the determination coefficient (R^2^), adjusted R^2^ (adj R^2^), F-value and p-value for each model without the interaction term. Additionally, we report the R^2^ change, F-value change and p-value of the model with the interaction between the disease effect and age. STN = sub-thalamic nucleus, L=left, R= right, * indicates a significant (p < 0.05) model - predictor.

| Map | Deep brain region | Model predictors p-value |  |  | R<sup>2</sup> | Adj R<sup>2</sup> | F-value (degree of freedom) | p-value | Interaction disease – age |  |  |
| --- | --- | --- | --- | --- | --- | --- | --- | --- | --- | --- | --- |
|  |  | Age | Disease presence | Disease duration |  |  |  |  | R<sup>2</sup> change | F-value change (degree of freedom) | p-value |
| χ | Caudate L | 0.001* | 0.910 | 0.579 | 0.321 | 0.283 | 8.355 (3,53) | <0.001* | 0.070 | 5.96 (1,52) | 0.185 |
|  | Caudate R | 0.007* | 0.402 | 0.101 | 0.279 | 0.211 | 6.007 (3,53) | 0.001* | 0.026 | 1.845 (1,52) | 0.180 |
|  | Putamen L | 0.097 | 0.323 | 0.009* | 0.284 | 0.228 | 6.977 (3,53) | <0.001* | <0.005 | 0.035 (1,52) | 0.853 |
|  | Putamen R | 0.084 | 0.162 | 0.008* | 0.319 | 0.267 | 7.932 (3,53) | <0.001* | 0.009 | 0.724 (1,52) | 0.399 |
|  | Pallidum L | <0.001* | 0.001* | <0.001* | 0.669 | 0.65 | 35.636 (3,53) | <0.001* | 0.001 | 0.200 (1,52) | 0.657 |
|  | Pallidum R | <0.001* | 0.001* | <0.001* | 0.661 | 0.642 | 34.465 (3,53) | <0.001* | 0.005 | 0.794 (1,52) | 0.377 |
|  | Thalamus L | 0.889 | 0.285 | 0.304 | 0.188 | 0.142 | 4.097 (3,53) | 0.110 | <0.005 | 0.026 (1,52) | 0.872 |
|  | Thalamus R | 0.225 | 0.289 | 0.504 | 0.130 | 0.125 | 3.542 (3,53) | 0.156 | 0.001 | 0.027 (1,52) | 0.870 |
|  | Substantia nigra L | 0.008* | 0.013* | <0.001* | 0.514 | 0.487 | 18.718 (3,53) | <0.001* | <0.005 | 0.001 (1,52) | 0.981 |
|  | Substantia nigra R | 0.005* | 0.003* | 0.002* | 0.549 | 0.523 | 21.487 (3,53) | <0.001* | 0.001 | 0.132 (1,52) | 0.718 |
|  | STN L | 0.208 | 0.136 | 0.085 | 0.210 | 0.165 | 4.692 (3,53) | 0.006* | 0.029 | 2.013 (1,52) | 0.162 |
|  | STN R | 0.056 | 0.044 | 0.044 | 0.348 | 0.298 | 9.275 (3,53) | <0.001* | 0.003 | 0.277 (1,52) | 0.601 |
|  | Red nucleus L | 0.942 | 0.043 | 0.278 | 0.297 | 0.257 | 7.459 (3,53) | <0.001* | 0.002 | 0.168 (1,52) | 0.684 |
|  | Red nucleus R | 0.044 | 0.172 | 0.533 | 0.343 | 0.293 | 8.530 (3,53) | <0.001* | 0.343 | 1.391 (1,52) | 0.244 |
|  | Cerebellar dentate L | 0.105 | 0.944 | 0.364 | 0.110 | 0.060 | 2.184 (3,53) | 0.101 | 0.006 | 0.324 (1,52) | 0.572 |
|  | Cerebellar dentate R | 0.256 | 0.303 | 0.845 | 0.157 | 0.109 | 3.295 (3,53) | 0.127 | 0.022 | 1.379 (1,52) | 0.246 |
| R2* | Caudate L | 0.050* | 0.184 | 0.765 | 0.295 | 0.256 | 7.410 (3,53) | <0.001* | 0.003 | 0.210 (1,52) | 0.649 |
|  | Caudate R | 0.034* | 0.272 | 0.679 | 0.288 | 0.247 | 7.133 (3,53) | <0.001* | 0.006 | 0.477 (1,52) | 0.493 |
|  | Putamen L | 0.063 | 0.773 | 0.517 | 0.162 | 0.115 | 3.426 (3,53) | 0.024* | 0.003 | 0.216 (1,52) | 0.644 |
|  | Putamen R | 0.060 | 0.872 | 0.646 | 0.163 | 0.116 | 3.450 (3,53) | 0.023* | 0.005 | 0.295 (1,52) | 0.589 |
|  | Pallidum L | 0.049* | 0.459 | 0.570 | 0.230 | 0.186 | 5.264 (3,53) | 0.003* | <0.005 | 0.012 (1,52) | 0.915 |
|  | Pallidum R | 0.041* | 0.522 | 0.636 | 0.221 | 0.177 | 5.010 (3,53) | 0.004* | <0.005 | 0.018 (1,52) | 0.895 |
|  | Thalamus L | 0.071 | 0.993 | 0.509 | 0.126 | 0.076 | 2.536 (3,53) | 0.067 | <0.005 | 0.029 (1,52) | 0.866 |
|  | Thalamus R | 0.145 | 0.598 | 0.760 | 0.131 | 0.082 | 2.667 (3,53) | 0.057 | 0.001 | 0.060 (1,52) | 0.807 |
|  | Substantia nigra L | 0.018 | 0.013 | <0.005 | 0.082 | 0.030 | 1.586 (3,53) | 0.204 | 0.008 | 0.433 (1,52) | 0.514 |
|  | Substantia nigra R | 0.228 | 0.719 | 0.710 | 0.090 | 0.038 | 1.741 (3,53) | 0.170 | 0.004 | 0.235 (1,52) | 0.630 |
|  | STN L | 0.075 | 0.996 | 0.424 | 0.128 | 0.079 | 2.592 (3,53) | 0.062 | <0.005 | 0.004 (1,52) | 0.953 |
|  | STN R | 0.100 | 0.883 | 0.493 | 0.095 | 0.044 | 1.855 (3,53) | 0.149 | <0.005 | 0.007 (1,52) | 0.934 |
|  | Red nucleus L | 0.259 | 0.997 | 0.564 | 0.058 | 0.005 | 1.091 (3,53) | 0.361 | 0.005 | 0.272 (1,52) | 0.604 |
|  | Red nucleus R | 0.232 | 0.892 | 0.510 | 0.055 | 0.002 | 1.033 (3,53) | 0.386 | 0.009 | 0.480 (1,52) | 0.491 |
|  | Cerebellar dentate L | 0.100 | 0.128 | 0.508 | 0.251 | 0.209 | 5.920 (3,53) | 0.100 | 0.009 | 0.630 (1,52) | 0.431 |
|  | Cerebellar dentate R | 0.124 | 0.487 | 0.805 | 0.158 | 0.110 | 3.319 (3,53) | 0.027 | 0.001 | 0.033 (1,52) | 0.856 |

There was not a statistically significant effect of disease on the relationship between age and the mean χ values within any of the structures as evidenced by increase in total variation explained (p>0.100) as reported on Table 2. Similar results were obtained for the model applied to the patients’ sub-groups.

R2* age and disease relationships in the deep brain nuclei were similar to those found for the QSM. The multiple regression model significantly (p<0.05) predicted the mean R2* in the caudate (left and right), putamen (left and right) and pallidum (left and right), as reported on Table 2. Age was a significant (p>0.05) predictor for the R2* values in the caudate and pallidum (see Table 2). The regression models applied on the healthy controls and the patients’ sub-groups showed significant age correlation in the same nuclei identified in the whole group analysis.

There was not a statistically significant effect of disease on the relationship between age and the mean R2* values within any of the structures as evidenced by increase in total variation explained (p>0.100) as reported on Table 2. Similar results were obtained within the FCDIIb and well-localised patient sub-groups.

## Discussion

To our knowledge, this is the first study using QSM to characterise epileptogenic lesions in humans. QSM was evaluated in a paediatric population of forty patients with drug-resistant focal epilepsy.

Our primary hypothesis was that *local* reductions in χ will be found in lesions driven primarily by alterations in their iron and calcium content. Consistent with our hypothesis, we demonstrated significant local χ decrease in MCD lesions, which was associated with reduced iron and myelin in several patients, and increased calcium and zinc in some cases. Measurements of across cortical depth in groups with well-localised and confirmed FCDIIb lesions both showed a reduction in χ at 2-3mm compared with homologous cortical regions. In two cases, the reduction in was associated with lower iron content that was linked with a pathological reduction in myelin observed by SRXRF and histopathological analyses, respectively.

Our second hypothesis was that *global* changes in brain iron in the deep grey matter nuclei might be present in children with poorly controlled focal epilepsy compared to healthy control subjects. While we found a positive correlation between age and both χ and R2* values in deep brain nuclei, consistent with previous reports (HALLGREN and SOURANDER, 1958; Ordidge et al., 1994; Yao et al., 2009; Ashraf et al., 2018; Ghassaban et al., 2019), there was also a positive correlation with disease duration.

### Alterations in cortical structure detected by QSM

The quantitative lesion profiling analysis showed consistent changes in for well-localised lesions and this was also seen in the FCDIIb sub-group. The analysis of with cortical depth (Figure 6) showed that lesions exhibited negative χ values across different sampling depths. In order to shed light on the tissue mineral substrate underlying these negative χ values, SRXRF was performed to identify and quantify alterations in multi-elemental concentrations. Increased calcium and zinc were observed in one FCDIIb sample. Both substances are diamagnetic and their presence is in agreement with hypo-intense QSM (and hyper-intense R2*) observed in the same sample. Previous studies on an animal model for epilepsy, revealed the presence of localized calcium deposits in diamagnetic regions delineated on QSM (Lafreniere *et al*., 1992; Aggarwal et al., 2018), however the alterations in zinc observed here, have not been previously described.

Dystrophic calcifications have been observed in a spectrum of neurological disorders, and are associated with abnormal mineral accumulation in areas of degenerated or necrotic tissue (Casanova and Araque, 2003). Therefore, reduced is likely to delineate lesions that have a high degree of tissue damage. High zinc levels have also been related to excitotoxicity in a variety of conditions, including ischemia, epilepsy, and brain trauma (Sensi et al., 2009, 2011). Zinc accumulation can cause both neuronal and glial death *in vitro* and *in vivo* (Aizenman et al., 2000; Hwang et al., 2008). Although the precise mechanisms of brain tissue mineralization are not currently understood, localized increase in intracellular calcium and zinc concentration have been associated with apoptotic or necrotic cell death and inflammatory processes (Casanova and Araque, 2003; Sensi et al., 2009, Liu et al., 2015*b*). Moreover, we found that, while the lesions showed uniform negative χ across sampling depths, homotopic regions had positive χ that peaked between 2-3.5mm depth. High resolution QSM maps acquired at 7T have shown that deeper layers of the cortex present increased χ values due to the presence of an iron-rich layer (Fukunaga et al., 2010; Marques et al., 2010). Our findings at 3T of increased χ values in non-lesional cortex, peaking at ~2.5mm are entirely consistent with a high iron layer corresponding to underlying myeloarchitecture. Although the myeloarchitecture changes across cortical regions, there is a consistent pattern of high myelination of deep cortical layers compared to superficial ones (Timmler and Simons, 2019). This finding is concordant with the few QSM studies that have evaluated whether iron is co-localised with myelin in deep cortical layers in healthy subjects (Deistung et al., 2013; Stüber et al., 2014).

This pattern of increased χ values at 2-3.5mm depth was absent in lesions both in the well-localised, and the histologically confirmed FCDIIb sub-group. Some FCD lesions sub-types have disrupted radial and tangential laminar structure of the cortex (Blumcke et al., 2017), that would be expected to alter typical myeloarchitecture, particularly in deep cortical layers and at the grey-white matter interface. Further, FCDIIb is associated with demyelination (Blumcke et al., 2017), consistent with the T2 FLAIR hyperintensity often seen radiologically (Blümcke et al., 2011). The absence of this peak in χ at 2-3.5mm cortical depth represents a potential signature of cortical dysplasia that could be useful for its identification and classification.

MCD lesions also have alterations in cytoarchitecture, with FCDIIb having dysmorphic neurons and balloon cells. The alterations in mineral content that might be associated with these abnormalities are largely unknown. SRXRF analysis of tissue samples of the both FCDIIb patients showed decreased iron in the brain tissue from the lesion. The iron decrease was spatially concordant with the myelin depletion highlighted by histological staining (see Figures 3 and 4), which could explain both the overall decrease in χ and the lack of the peak in χ values in the lesional profile.

Further evidence for this finding includes the lack of a decrease in iron in the control tissue. We found that when including subjects with poorly localised focal epilepsy (for example discrepant seizure semiology and MRI lesion location) we did not find significant χ profile differences between FCD lesions and homotopic regions (see Figure 6: top row). This might suggest that the reduced values are specific to the true epileptogenic lesion. The results found here motivate future studies, designed and powered to evaluate the potential to use this signature of MRI χ changes for lesion detection and stratification.

SRXRF elemental mapping on the specimen with glioneuronal tumour revealed increased calcium and zinc in tumour tissue. The presence of these diamagnetic substances is consistent with hypo-intensity on QSM and hyper-intensity on R2*. The pathophysiology underlying the mineralization of this brain tumour is not currently understood (Deistung et al., 2013; Ciraci et al., 2017; Eskreis-Winkler et al., 2017). The tumour specimen also showed the presence of increased iron that could be due to bleeding. Notably, data was obtained from only one patient and more studies are required to determine trace elemental concentrations in such brain tumours (Cilliers et al., 2020).

### Age effect in deep brain nuclei

In agreement with previous studies carried out in paediatric population (Li et al., 2014; Darki et al., 2016; Zhang et al., 2018; Peterson et al., 2019), a significant linear correlation was observed between age and χ measured in the caudate, globus pallidus, substantia nigra, sub-thalamic nucleus and red nucleus for the whole cohort of patients and healthy controls. Moreover, we found a significant linear dependency between age and the mean R2* in the caudate and globus pallidus. These nuclei are known to have the greatest age-related iron accumulation of all the deep brain structures investigated in this study, leading to increased χ and R2* values, (Li et al., 2014; Zhang et al., 2018).

Other structures such as the putamen, thalamus and cerebellar dentate are also known to accumulate iron, however at a slower rate or resulting in a lower mean susceptibility value, and, therefore, correlations between χ / R2* and age in these regions did not reach statistical significance given the relatively small group size (n=40) and age range (2-21). It should also be noted that the spatial location of focal epilepsy varied within this cohort and, therefore, more spatially localised changes in regions such as the thalamus could be present in larger more homogenous cohorts reflecting volumetric and functional connectivity studies (Centeno et al., 2014; He et al., 2017). We found potential evidence increases in brain iron due to epilepsy duration that could reflect accelerated brain-aging in this paediatric cohort. In this patient group seizure onset is typically early in life and therefore age and duration of epilepsy are highly correlated. Never-the-less our model did suggest that duration of epilepsy did explain significant additional differences in several deep brain nuclei. This interesting finding should be confirmed in a group with a wider range of onsets and ages.

### Visual assessment

Based on the multiple comparison statistical tests, the QSM and R2* maps provided worse FCD lesion conspicuity compared to FLAIR and T1w images. At the individual level, the lesion contrast on QSM and R2* was enhanced in respectively 5% and 7.5% of patients; therefore, these maps are most likely to provide information in combination with FLAIR to detect and characterise MCD. QSM provided very high contrast in a small but significant number of patients which was related to calcium and zinc increases. The sensitivity to calcification may be useful for targeting in cases with widespread abnormalities such as tuberos sclerosis. Future studies are needed to validate the radiological value of maps in larger cohorts including in MRI-negative patients.

### Limitations and outlook

The clinical trend is to perform surgical resection on patients with clearly defined epileptogenic regions and seizures that affect both quality of life and development. It is therefore crucial to non-invasively characterise focal lesions in order to improve surgical planning, seizure freedom and neuro-developmental outcome (Adler et al., 2017; Choi et al., 2018). This study is the first investigation using QSM in epileptic lesions to reveal alterations in iron content that appear to be related to myeloarchitecture associated with MCD. An observed MRI abnormalities confined to the epileptogenic zone (presumed n=4, confirmed n=21) was reduced and the absence of a peak in χ at a cortical depth of ~2.5 mm. The consistency of χ changes across different cortical regions in FCDIIb lesions indicates that these maps could be useful for the characterisation of those lesions and could be integrated into algorithms for automated lesion detection (Adler et al., 2017; Lorio et al., 2020). Future studies are needed to validate cortical QSM profiles for automated lesion detection in larger cohorts.

In 5% of patients, the lesions were visually more conspicuous on QSM with respect to FLAIR and T1w images. Some lesions with negative χ were identified and further analysed with SRXRF in two patients with FCDIIb and one patient with a glioneuronal tumour showing calcium and also zinc increases. The clinical relevance of these changes in tissue composition is not clear. However, in tuberous sclerosis, which has histopathological similarities to FCDIIb, calcification has been reported to be dynamic and associated with epileptogenicity (GALLAGHER et al., 2010). The high zinc levels detected non-invasively with QSM may be related to neurotoxicity and gliotic changes. The presence of extracellular Zn^2+^ is known to alter neuronal excitability and therefore facilitate and modulate epileptiform activity in the hippocampus (Coulter, 2000; Carver et al., 2016), while its role in epileptogenic cortical abnormalities remains largely unknown.

QSM is a rapidly developing field of MRI with increasingly sophisticated acquisition and, in particular, complex post-processing algorithms. The magnetic susceptibility is estimated from the MRI signal phase, which is affected by the contributions from local microscopic tissue composition and macroscopic background fields which are removed via post-processing. QSM is also sensitive to acquisition parameters (particularly TE) (Biondetti et al., 2020) and tissue orientation with respect to the main magnetic field (Schweser et al., 2011; Wharton and Bowtell, 2012, 2015). Despite these limitations we found consistent differences in susceptibility values in the cortex. This is a unique finding because QSM has habitually been applied to iron-rich deep brain regions (where we also found age-related susceptibility increases). In this study QSM was performed at 3T, demonstrating clinical relevance, however phase-based measurements, such as QSM, benefit from increases in contrast and resolution at 7T, therefore this study strongly motivates further application of QSM to study patients with epilepsy at 7T.

## Data Availability

This data involves patients, data can be made available in anonymous format please contact David Carmichael.

## Acknowledgements

This research was supported by the NIHR Great Ormond Street Hospital Biomedical Research Centre. The views expressed are those of the author(s) and not necessarily those of the NHS, the NIHR or the Department of Health. We thank the Diamond Light Source for access to the I18 beamline for synchrotron-radiation X-ray fluorescence elemental mapping.

## Funding

This research was funded by the Henry Smith Charity and Action Medical Research (GN2214). David Carmichael and Sara Lorio were supported by the King’s College London Wellcome/EPSRC Centre for Medical Engineering [WT 203148/Z/16/Z]. SA received funding from the Rosetrees Trust. TSJ receives funding from Great Ormond Street Children’s Charity, The Brain Tumour Charity, Children with Cancer UK, Cancer Research UK, NIHR and the Olivia Hodson Cancer Fund. KS is supported by a European Research Council Consolidator Grant [DiSCo MRI SFN – 770939].

## Competing interests

None of the authors has any conflict of interest to disclose.

## Supplementary material

### Healthy controls

17 healthy controls (mean age = 15 ± 3 years, range= 8-21 years, female=10) were scanned for this research study at Great Ormond Street Hospital on a 3T whole-body MRI system (Magnetom Prisma, Siemens Medical Systems, Germany), using a 64-channel receive head coil and body coil for transmission. Three-dimensional structural T1w images were acquired using MPRAGE, FLAIR, and a multi-echo T2*-weighted sequence. The acquisition parameters were the same as the ones used for the patients’ data acquisition.

## Histology staining

Formalin-Fixed-Paraffin-Embedded (FFPE) tissue blocks were obtained from Great Ormond Street Hospital (GOSH) for Haematoxylin and Eosin staining (H&E), Luxol Fast Blue (LFB), Neuropathology special stain, Immunohistochemistry (IHC) and slide scanning.

### H&E staining

Sections were cut at 5μm thickness and stained on Leica ST5010/CV5030 automated workstation. The automated machine incubated the sections in 2 changes of xylene (Genta Medical UN1307), followed by rehydration in graded alcohols (Genta Medical UN1170) and a wash step in distilled water. Next, the sections were stained with Harris Haematoxylin (Leica Surgipath; Ref 3801560E) for 6 minutes 30 seconds exactly, then washed in distilled water and briefly differentiated in 1% acid alcohol for 10 seconds to remove excess haematoxylin. Subsequently, the sections were blued in distilled water followed by counterstaining in 1% Eosin (Leica Surgipath; Ref 3801590E) for approximately 4 minutes 30 seconds. Afterward, the sections were washed in distilled water, dehydrated in graded alcohols and cleared in 2 changes of xylene in preparation for mounting. The sections were mounted with Pertex (Histolab; Ref 00801) mounting media for light microscopy.

### Luxol Fast Blue (LFB)

Sections were cut at 14 μm on super frost adhesive slides, baked overnight in 37 °C oven and in 60°C oven for 1 hour the following morning. Slides were de-waxed in xylene, rehydrated in graded alcohols and placed in a staining dish containing filtered luxol fast blue in acidified methanol (Atom Scientific; Ref RRSP248-G, Lot 116149). The slides were immediately transferred into a water bath pre-heated to 60°C and left to incubate for 2 hours. Afterward, the sections were briefly rinsed with denatured 70% ethanol (Atom Scientific; Ref RRSP242-G, Lot 116642) and distilled water before differentiating in 0.05% lithium carbonate solution (Atom Scientific; Ref RRSP252-G, Lot 113742) until the grey and white matter can be distinguished. Subsequently, the slides were rinsed in 95% alcohol and water followed by counterstaining with cresyl violet 0.5% aqueous solution (Atom Scientific; Ref RRSP251-G, Lot 116921) for 10-12 minutes. Briefly, the sections were washed in water and differentiated in a solution of 1% acetic acid in 95% denatured ethanol (Atom Scientific; Ref RRSP260-G, Lot 113792) for up to 4 seconds. The sections were rapidly dehydrated through graded alcohols, cleared in 2 changes of xylene and mounted with pertex mounting media as previously described.

### Immuno-histochemistry (IHC)

IHC staining was performed on a Leica Bond-Max auto-stainer (Leica Bio systems, Melbourne, Australia) and the Leica Bond Polymer Refine Detection kit (Ref. DS9800) was used to visualise bound antibody. Neuron specific protein expression was detected using anti-NeuN (clone A60), mouse monoclonal primary antibody catalogue no. MAB377 (Merck Millipore, Billerica, MA, USA), with an optimized dilution of 1:500. To validate IHC staining, a surgical brain tissue with known expression of the target protein was used as positive control and Tissue Micro Array (TMA) was used as negative control by omitting the application of the primary antibody. On the Leica Bond-Max machine, the sections were de-waxed with xylene and rehydrated with 99% Industrial Denatured Alcohol (IDA). Antigen retrieval was achieved by Heat Induced Epitope Retrieval (HIER), Leica Epitope Retrieval protocol 2 (ER2) for 30 minutes, pH 9, Bond-max protocol F. Peroxidase block was applied for 5 minutes with Bond polymer refine kit (Ref. DS9800) followed by the application of the primary antibody for 1 hour. The sections were subsequently incubated with post primary (Bond polymer refine kit; Ref. DS9800) for 8 minutes and HRP labelled polymer (Bond polymer refine kit; Ref. DS9800). The application of 3, 3-Diaminobenzidine (DAB) (Bond polymer refine kit; Ref. DS9800) chromogen solution to the sections forms a brown precipitate at positively expressed antigen sites. The tissue sections were counterstained with haematoxylin (Bond polymer refine kit; Ref. DS9800) and mounted for light microscopy as previously described. The test tissues were cut at 5μm & controls at 3 μm thickness and mounted on a Leica Surgipath X-tra adhesive slides.

### Slide scanning

The stained sections were scanned on the Leica Aperio CS2 Scanner (S/N 5872) at 40x objective.

## Notes

### Competing Interest Statement

The authors have declared no competing interest.

### Clinical Trial

This is a basic science study on human participants

### Author Declarations

UK National Research Ethics Service Project Title:Using MRI tissue parameter maps to detect and delineate FCD IRAS Project ID: 148540 Project Type:Basic science study involving procedures with human participants

